# The Diabetes Epidemic in Bangladesh: Trends and Forecasts (2000 to 2050)

**DOI:** 10.64898/2025.12.25.25342994

**Authors:** Jannatul Nayma Eshita, Abu Bakar Al Mizan, Istekab Bin Yousuf, Tazeen Tarannum Sadia, Sidratul Muntaha, Sumaia Alam, Abdullah Al Rafi, Debabrata Mallick

**Affiliations:** Department of Biochemistry and Biotechnology, University of Science and Technology Chittagong (USTC), Chattogram-4202, Bangladesh; Department of Computer Science and Engineering, University of Science and Technology Chittagong (USTC), Chattogram-4202, Bangladesh

## Abstract

To examine long-term trends in the prevalence and burden of diabetes among adults in Bangladesh from 2000 to 2050 using historical estimates and future projections.A descriptive epidemiological trend analysis using secondary data from the International Diabetes Federation (IDF) Diabetes Atlas (2000, 2011, 2024, and 2050 projections) and prevalence estimates calibrated with the Bangladesh Demographic and Health Survey (BDHS) 2017– 2018.Bangladesh, South-East Asia Region. Adults aged 20–79 years included in IDF diabetes estimates; additional population-level prevalence information drawn from BDHS. Number of adults with diabetes and age-standardised diabetes prevalence. The number of adults (20–79 years) with diabetes increased from 1.8 million in 2000 to 13.9 million in 2024, and is projected to reach 23.1 million by 2050. Age-standardised prevalence rose from 10.5% in 2011 to 13.2% in 2024, with a further increase to 15.4% projected for 2050. In 2024, 39.1% of adults with diabetes remained undiagnosed. Impaired fasting glucose and impaired glucose tolerance affected 19.1 million and 14.5 million adults respectively in 2024, with substantial projected increases by 2050. Bangladesh ranks second highest in the South-East Asia Region in both prevalence and total number of adults with diabetes, following India. Diabetes-related mortality accounted for 31,619 deaths in 2024, and health expenditure reached USD 1.03 billion, increasing further in future projections. Bangladesh faces a rapidly escalating diabetes epidemic, marked by sharp increases in prevalence, undiagnosed cases, metabolic risk conditions, and healthcare expenditure. Without immediate public health interventions particularly lifestyle modification programmes, expanded screening, and strengthened surveillance the diabetes burden is expected to rise substantially by 2050.

**STRENGTHS AND LIMITATIONS OF THIS STUDY:**

⇒ We examined long-term changes in the diabetes burden among Bangladeshi adults from 2000 to 2024 and projections up to 2050.
⇒ Estimates were derived from nationally representative and internationally validated data sources, including BDHS and IDF.
⇒ This study uniquely combines historical trends with future forecasts to highlight the evolving epidemiology of diabetes.
⇒ All data were generated following standardize and WHO-aligned assessment methods.
⇒ Limited availability of continuous annual data and missing risk factor variables restricted deeper analytical exploration.

## INTRODUCTION

Diabetes mellitus has emerged as one of the most pressing non-communicable diseases (NCDs) globally, with disproportionate growth in low- and middle-income countries **[1,2]**. Bangladesh, a densely populated South Asian nation undergoing rapid demographic and epidemiological transition, is experiencing an unprecedented rise in diabetes prevalence.

According to the International Diabetes Federation (IDF), Bangladesh is one of the seven countries in the South-East Asia Region and currently holds the second highest diabetes prevalence (20–79 years) within the region **[3]**. Over the past two decades, the number of adults aged 20–79 years living with diabetes has increased from 1.8 million in 2000 to 8.4 million in 2011, reaching 13.9 million in 2024, with projections indicating a further escalation to 23.1 million by 2050 **[3,4]**. These trends reflect a complex interplay of population growth, urbanization, sedentary lifestyles, dietary shifts, and limited screening and early diagnostic capacity **[5–7]**.

The accelerating diabetes epidemic poses substantial challenges for Bangladesh’s health system, including increased healthcare costs, heightened cardiovascular risk, and long-term disability among working-age adults **[6,8]**. Although several national surveys and demographic health studies such as the Bangladesh Demographic and Health Survey (BDHS) 2017–18, conducted by the National Institute of Population Research and Training (NIPORT) and ICF have documented rising NCD prevalence, a comprehensive synthesis of long-term trends and future forecasts for diabetes is lacking **[5,9]**.

Understanding these patterns is essential for designing targeted public health interventions, strengthening surveillance, and informing policy decisions aligned with national NCD action plans.

This study aims to examine the historical trend of diabetes in Bangladesh from 2000 to 2024 and forecast the burden up to 2050 using available epidemiological estimates. By providing a detailed assessment of past growth and projected trajectories, this analysis seeks to highlight the magnitude of the challenge and the urgency for prevention-focused, multisectoral responses **[3,10]**.

## METHODS

### Data source

We used secondary data from the International Diabetes Federation (IDF) Diabetes Atlas and the Bangladesh Demographic and Health Survey (BDHS) 2017–18 to estimate the prevalence and number of adults (20–79 years) with diabetes in Bangladesh. The IDF Diabetes Atlas provides standardized estimates of diabetes prevalence and projections for all countries, while the BDHS 2017–18 collected nationally representative data on population health, risk factors, and demographic characteristics. Additional contextual information was obtained from previous BDHS waves and the WHO STEPS survey 2018 to assess trends and risk factors **[3–7]**.

### Study population

The study focused on adults aged 20–79 years in Bangladesh. IDF data provided population-level estimates for 2000, 2011, 2024, and projected values for 2050. No individual-level data were used; the analysis relied entirely on aggregated and de-identified datasets **[3,4,5]**.

### Outcome variable

The primary outcome was the estimated prevalence and total number of adults with diabetes (20–79 years). Diabetes prevalence was derived from IDF standardized definitions, including fasting plasma glucose ≥7.0 mmol/L or self-reported use of glucose-lowering medication, as reported in national surveys **[3,5]**.

### Covariates and contextual variables

To assess demographic and lifestyle influences, we included information on age distribution, sex, urban versus rural residence, body mass index (BMI), overweight/obesity prevalence, and regional differences in diabetes burden. Population growth, urbanization, dietary transition, and physical activity trends were also considered when interpreting projected increases **[5–7,9–11]**.

### Analytical approach

Historical trends from 2000 to 2024 were analyzed by calculating absolute and relative changes in diabetes prevalence and total affected adults. The 2050 projections were interpreted to estimate future burden, considering demographic growth, population aging, and risk factor patterns modeled by the IDF **[3,4]**. Bangladesh’s ranking within the IDF South-East Asia Region was assessed descriptively to provide regional context, comparing prevalence and total number of adults with diabetes to neighboring countries such as India and Pakistan **[3]**. A narrative synthesis of demographic transitions, lifestyle changes, and national screening coverage contextualized numeric trends.

### Ethical considerations

All data were aggregated, publicly available, and de-identified; no individual-level data were accessed. Ethical approval was therefore not required **[5,12]**

## RESULTS

### Diabetes Burden and Prevalence

Table 1. shows the estimated number of adults (20–79 years) with diabetes in Bangladesh and India from 2000 to 2050. The number of people with diabetes in Bangladesh increased from 1,759.7 thousand in 2000 to 8,405.6 thousand in 2011, 13,877.4 thousand in 2024, and is projected to reach 23,071.0 thousand in 2050. Similarly, India experienced a rise from 32,674.4 thousand in 2000 to 61,258.4 thousand in 2011, 89,826.9 thousand in 2024, and a projected 156,740.7 thousand in 2050. This demonstrates a sharp upward trend in diabetes burden in both countries, with Bangladesh showing a relatively faster proportional increase over time.

**Table 1.** Adults (20–79 years) With Diabetes in Bangladesh and India (2000–2050)

| Year | Bangladesh (1,000s) | India (1,000s) |
| --- | --- | --- |
| 2000 | 1,759.70 | 32,674.40 |
| 2011 | 8,405.60 | 61,258.40 |
| 2024 | 13,877.40 | 61,258.40 |
| 2025 | 23,071.00 | 156,740.70 |

The age-standardised prevalence of diabetes is presented in **Table 2**. Bangladesh had a prevalence of 10.5% in 2011, which increased to 13.2% in 2024 and is projected to reach 15.4% in 2050. India’s prevalence was 9.0% in 2011, rising to 10.5% in 2024 and 12.8% in 2050. These figures indicate a consistent upward trend in prevalence, reflecting increasing disease burden beyond population growth alone. **Table 3** presents the proportion and number of undiagnosed diabetes cases in 2024. In Bangladesh, 39.1% of adults with diabetes remained undiagnosed, corresponding to 5,426.1 thousand individuals, while in India, 43% remained undiagnosed, totaling 38,598.6 thousand people. Data for other years were not available, highlighting a critical gap in diagnosis and early detection.

**Table 2.** Age-Standardised Prevalence of Diabetes (%)

| Year | Bangladesh | India |
| --- | --- | --- |
| 2000 |  |  |
| 2011 | 10.5 | 9 |
| 2024 | 13.2 | 10.5 |
| 2025 | 15.4 | 12.8 |

**Table 3.** Proportion and Number of Undiagnosed Diabetes.

| Year | Bangladesh | India |
| --- | --- | --- |
| 2024 | 1,031.80 | 9,834.40 |
| 2050 | 1,440.40 | 15,265.10 |

Impaired fasting glucose (IFG) trends are shown in **Table 4**. In 2024, 19,117.2 thousand adults in Bangladesh and 107,751.1 thousand in India had IFG, representing 17.1% and 11.7% of the adult population, respectively. By 2050, these numbers are projected to increase to 28,526.2 thousand (19.2%) in Bangladesh and 148,962.3 thousand (12.2%) in India, indicating rising prediabetes prevalence.

**Table 4.** Impaired Fasting Glucose (IFG) in Adults (20–79 years)

| Year | Bangladesh(1,000s) | India(1,000s) | Bangladesh % | India% |
| --- | --- | --- | --- | --- |
| 2000 |  |  |  |  |
| 2011 |  |  |  |  |
| 2024 | 19,117.20 | 107,751.10 | 17.1 | 11.7 |
| 2025 | 28526.2 | 148,962.30 | 19.2 | 12.2 |

Impaired glucose tolerance (IGT) estimates are presented in **Table 5**. In 2011, Bangladesh had 2,141.1 thousand adults (2.8%) with IGT, increasing to 14,536.3 thousand (13.5%) in 2024 and 20,535.4 thousand (13.8%) in 2050. India shows a similar trend, with 20,467.5 thousand (3.0%) in 2011, 127,292.1 thousand (13.9%) in 2024, and 179,285.8 thousand (14.8%) in 2050.

**Table 5.**
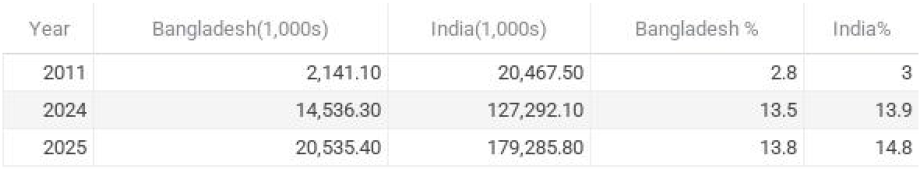
Impaired Glucose Tolerance (IGT) in Adults (20–79 years)

Mortality attributable to diabetes is reported in **Figure 1**. In 2011, Bangladesh experienced 144,443 deaths due to diabetes, which is projected to decrease to 31,619.5 in 2024. India reported 983,203 deaths in 2011 and 334,922.2 in 2024. The percentage of deaths among adults aged 20–79 years in 2024 was 5.6% in Bangladesh and 5.0% in India.

**Figure 1.**
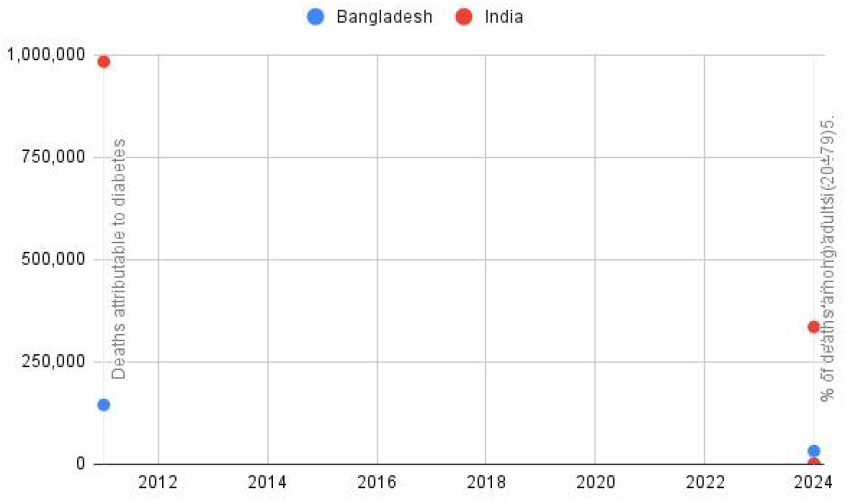
Mortality Attributable to Diabetes (20–79 years)

Hyperglycaemia in pregnancy (HIP) estimates are presented in **Table 7**. In 2024, 697,071.3 live births in Bangladesh and 6,272,667.4 in India were affected by HIP. The prevalence of gestational diabetes mellitus (GDM) was 18.5% in Bangladesh and 26.1% in India. Additionally, other diabetes first detected in pregnancy and diabetes detected prior to pregnancy are reported in the table.

**Table 7.** Hyperglycaemia in Pregnancy (20–49 years)

| Year | Indicator | Bangladesh | India |
| --- | --- | --- | --- |
| 2024 | Live births affected by HIP | 697,071.30 | 6,272,667.40 |
| 2024 | Prevalence of GDM (%) | 18.5 | 26.1 |
| 2024 | Other diabetes first detected in pregnancy | 74,159.50 | 217,165.30 |
| 2024 | Diabetes detected prior to pregnancy | 115,506.80 | 288,222.90 |

**Table 8(A).** Diabetes-Related Health Expenditure-Total Expenditure (USD million)

| Year | Bangladesh% | India% | Bangladesh (1,000s) | India(1,000s) |
| --- | --- | --- | --- | --- |
| 2000 |  |  |  |  |
| 2011 |  |  |  |  |
| 2024 | 39.1 | 43 | 5,426.10 | 38,598.60 |
| 2025 |  |  |  |  |

**Table 8(B).** Per-Person Expenditure (USD)

| Year | Bangladesh | India |
| --- | --- | --- |
| 2011 | 27 | 68 |
| 2024 | 74.4 | 109.5 |
| 2050 | 62.4 | 97.4 |

Diabetes-related health expenditures are shown in Table 8. Total expenditure in 2024 was 1,031.8 million USD in Bangladesh and 9,834.4 million USD in India, increasing to 1,440.4 million USD and 15,265.1 million USD in 2050, respectively. Per-person expenditure also increased over time, from 27 USD in 2011 to 74.4 USD in 2024 in Bangladesh, and from 68 USD to 109.5 USD in India.

**Type 1** diabetes estimates are presented in Table 9. In 2024, Bangladesh had 25,520 individuals with type 1 diabetes across all ages, including 7,425.9 aged 0–19 years. India had 940,840 people with type 1 diabetes overall, including 300,788.4 aged 0–19 years. Demographic indicators are summarized in **Table 10**. The adult population (20–79 years) in Bangladesh increased from 83,797 thousand in 2000 to 147,517 thousand in 2050, while India’s adult population increased from 567,714 thousand to 1,205,759.3 thousand over the same period. Finally, **Table 11** provides a South Asia regional context, including Pakistan. The number of adults with diabetes in 2050 is projected to be 23,071 thousand in Bangladesh, 156,741 thousand in India, and 70,189 thousand in Pakistan. Age-standardised prevalence in 2050 is estimated at 15%, 13%, and 34% in Bangladesh, India, and Pakistan, respectively, highlighting regional disparities in diabetes burden.

**Table 9.**
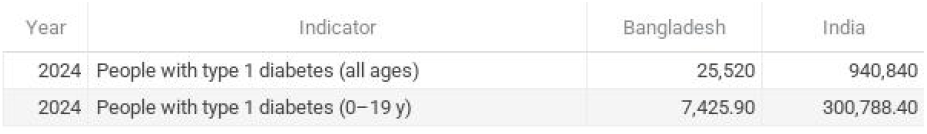
Type 1 Diabetes Estimates (All Ages and 0–19 years)

**Table 10.** Demographic Indicators (Adult Population 20–79 years)

| Year | Bangladesh (1,000s) | India (1,000s) |
| --- | --- | --- |
| 2000 | 83,797 | 567,714 |
| 2011 | 87,708.10 | 737,003.30 |
| 2024 | 113,054.50 | 947,373.60 |
| 2050 | 147,517 | 1,205,759.30 |

**Table 11.** South Asia regional context.

People with diabetes, in 1,000s
| Year | Bangladesh | India | Pakistan |
| --- | --- | --- | --- |
| 2000 | 1,760 | 32,674 | 8,790 |
| 2011 | 8,406 | 61,258 | 6,349 |
| 2024 | 13,877 | 89,827 | 34,532 |
| 2050 | 23,071 | 156,741 | 70,189 |

| Year | Bangladesh | India | Pakistan |
| --- | --- | --- | --- |
| 2000 | 0 | 0 | 12 |
| 2011 | 11 | 9 | 8 |
| 2024 | 13 | 11 | 31 |
| 2050 | 15 | 13 | 34 |

## Discussion

This study presents a comprehensive assessment of the diabetes burden in Bangladesh and India from 2000 to 2050, highlighting both historical trends and projected trajectories. Our findings indicate a rapid and substantial increase in the number of adults living with diabetes in both countries, with Bangladesh showing a particularly steep proportional rise despite having a smaller absolute population compared with India **(Table 1)**. The age-standardised prevalence of diabetes also demonstrated an upward trend in both countries, rising from 10.5% in 2011 to 15.4% in 2050 in Bangladesh, and from 9.0% to 12.8% in India **(Table 2)**. These increases reflect the combined effects of population growth, urbanization, ageing, and shifts in lifestyle and dietary patterns.

The high proportion of undiagnosed diabetes cases, estimated at 39.1% in Bangladesh and 43% in India in 2024 **(Table 3)**, underscores a significant gap in early detection and screening. Delayed diagnosis may contribute to higher complication rates, including cardiovascular diseases, renal dysfunction, and other diabetes-related comorbidities.

The rising prevalence of impaired fasting glucose (IFG) and impaired glucose tolerance (IGT) **(Tables 4 and 5)** further suggests a growing population at risk for developing type 2 diabetes, highlighting the need for targeted prevention strategies and lifestyle interventions.

Mortality attributable to diabetes, although showing some fluctuations, remains substantial. In 2024, diabetes contributed to 5.6% and 5.0% of adult deaths in Bangladesh and India, respectively **(Table 6)**, indicating a persistent public health burden. The increasing prevalence of hyperglycaemia in pregnancy (HIP) in both countries **(Table 7)** is also concerning, given its implications for maternal and neonatal health and the future risk of type 2 diabetes among offspring.

Economic implications of the diabetes epidemic are considerable. Total health expenditure attributable to diabetes is projected to rise to 1,440.4 million USD in Bangladesh and 15,265.1 million USD in India by 2050 **(Table 8)**. This underscores the financial strain on national healthcare systems and emphasizes the need for cost-effective prevention, early detection, and management strategies.

Type 1 diabetes, although less prevalent than type 2 diabetes, remains a critical concern, especially in children and adolescents **(Table 9)**. Timely diagnosis and lifelong management are essential to reduce morbidity and improve quality of life. Demographic shifts, particularly the rapid increase in the adult population **(Table 10)**, are likely to amplify the overall diabetes burden and demand for healthcare resources. Regional comparisons, including Pakistan, reveal notable disparities in both prevalence and projections **(Table 11)**, emphasizing the importance of context-specific public health interventions.

Overall, these findings highlight the urgent need for multisectoral strategies to address the rising diabetes burden in South Asia. Efforts should include strengthening primary care services, implementing national screening programs, promoting healthy diets and physical activity, and improving public awareness. Policymakers must prioritize resource allocation for both prevention and treatment to mitigate the long-term health, social, and economic impacts of diabetes.

### Strengths and Limitations

This study’s strengths include the use of internationally recognized datasets, standardized age-adjusted estimates, and comprehensive projections for both historical and future periods. However, limitations include the lack of intermediate yearly data for more precise trend modeling, unavailability of country-specific longitudinal data, and potential underestimation of undiagnosed diabetes cases.

## Conclusion

The diabetes burden in Bangladesh is rising rapidly, both in absolute numbers and age-standardized prevalence, with projections indicating a continued increase through 2050. A substantial proportion of adults remain undiagnosed, and a growing number of individuals exhibit impaired glucose regulation, highlighting a large population at risk for type 2 diabetes. These trends, coupled with rising diabetes-related mortality, pregnancy complications, and healthcare expenditures, underscore the urgent need for comprehensive public health strategies. Effective interventions should prioritize early detection, lifestyle modification, health education, and strengthened healthcare infrastructure. Policymakers must adopt multisectoral approaches that integrate prevention, treatment, and monitoring to mitigate the long-term health and economic impacts of diabetes in Bangladesh. Timely action is essential to curb the epidemic and reduce its burden on individuals, families, and the national health system.

## Data Availability

All data produced are available online at https://diabetesatlas.org/

